# Urinary miRNA Profiles with Machine Learning for Noninvasive Detection and Prognosis of Urological Malignancies

**DOI:** 10.1101/2025.10.11.25336309

**Authors:** Yuto Baba, Shun Iwasa, Yota Yasumizu, Toshikazu Takeda, Kazuhiro Matsumoto, Shinya Morita, Akira Cortal, Hiroki Yamaguchi, Yoriko Ando, Motoki Mikami, Mika Mizunuma, Yuki Ichikawa, Yasutaka Kato, Takeo Kosaka, Nobuyuki Tanaka, Mototsugu Oya

## Abstract

The need for biomarkers that can noninvasively determine and stratify cancer risk is emerging. MicroRNAs (miRNAs) are stable in urinary exosomes, allowing noninvasive detection of urinary tract cancers through the use of liquid biopsies. Herein, we proposed a novel diagnostic framework based on a urinary miRNA profile, enabling urological cancer screening and prognosis prediction. In total, 419 urine samples were prospectively collected and comprehensively sequenced, comprising three urological cancer cohorts—renal cell carcinoma (RCC, n=96), prostate cancer (PCa, n=153), and urothelial carcinoma (UC, n=131)—while healthy subjects were recruited as counterparts. Machine learning algorithms could distinguish patients with RCC, PCa, or UC from healthy individuals with high discriminatory performance for all three cancers, with AUCs of 0.92 for RCC, 0.92 for PCa, and 0.96 for UC. Furthermore, we developed urinary miRNA-based prognostic models using a panel of up to five urinary miRNAs for each cancer type, revealing time-dependent AUCs of recurrence-free survival ranging from 0.74 to 0.89 across all three cancer types at 48 months of follow-up, demonstrating high discriminatory power for long-term recurrence. Collectively, our findings support the potential use of primary miRNA signatures as a comprehensive, noninvasive tool for both diagnosis of and risk stratification for urological malignancies, positioning our urine-based assays as promising tools for future practice.

## INTRODUCTION

Genitourinary malignancies, including those of the kidney, prostate, and urothelium, remain among the most prevalent diseases worldwide^1,2^. Despite recent advances in high-resolution imaging and molecular diagnostics, current screening and surveillance methods suffer from several limitations, including invasiveness, poor specificity, and limited sensitivity, especially in early-stage disease^3,4^. For example, prostate-specific antigen testing is widely used for prostate cancer (PCa) screening; rather, blood sampling is invasive. For urothelial carcinoma (UC), cystoscopy remains the gold standard for diagnosis, but the invasiveness and cost of cystoscopy place a significant burden on both patients and health care systems^5^. Unfortunately, incidental findings are common in kidney cancer, especially small renal tumors, and there is a lack of robust biomarkers, both invasive and non-invasive, complicating early diagnosis and risk stratification^6^. Together, these facts highlight the unmet medical needs for potential biomarkers that can noninvasively detect cancer in the early stage, stratify future risk, and connect patients to appropriate medical care^7^.

MicroRNAs (miRNAs) are small noncoding RNAs of approximately 22 nucleotides that regulate gene expression at the posttranscriptional level and have recently emerged as candidates for liquid biopsy^8^. Normally, miRNAs are quickly degraded in their free state, but because they are small molecules, they exist in extracellular vehicles (EVs) such as exosomes and are retained in various bodily fluids^9,10^. In other words, miRNAs in exosomes are protected not only in the blood but also in urine, allowing them to be excreted from the body via urine with high stability^11,12^.

Therefore, in recent years, research investigating urinary EV-derived miRNAs as biomarkers for cancer detection, staging, and prognosis prediction has attracted attention in both basic and clinical settings^13^. In this regard, due to the direct contact between urine and the genitourinary system^8^, we hypothesized that tumor-derived EVs may be directly excreted into the urine in kidney, prostate, and urothelial malignancies; thus, they may be ideal biomarker candidates for the noninvasive detection of urinary tract cancers in liquid biopsies.

In this study, we proposed a novel diagnostic framework enabling urological cancer screening and prognosis prediction based on urine-derived EVs. We focused on the miRNA profile in EVs in urine and prospectively developed and evaluated machine learning algorithms for the detection of renal cell carcinoma (RCC), PCa, and UC. Concurrently, we investigated a miRNA-based prognostic model based on recurrence-free survival. Our findings may support the potential utility of urinary miRNA signatures as a comprehensive, noninvasive tool for both diagnosis and risk stratification in patients with urological malignancies.

## METHODS

### Ethics and Participants

All procedures were performed in compliance with the 1964 Helsinki Declaration and its later amendments. Human sample studies were performed with the approval of the Research Ethics Committee of Keio University (Approval No. 20180352) and Craif, Inc. (No. IEF-20220516). Participation in the study was optional, and written informed consent procedures were applied; patients with RCC, PCa, and UC were prospectively enrolled between 2019 and 2022 at Keio University Hospital. Patients were eligible if they were aged 20 years or older and had a confirmed diagnosis based on pathological examination of tumor specimens. Patients were classified according to the American Joint Committee on Cancer staging system (8th edition). Individuals with multiple primary cancers were excluded. For the control cohort, urine samples were collected from healthy volunteers aged 20 years or older without a history of cancer within the preceding five years and without active urological symptoms.

### Urine Collection and RNA Extraction

Urine samples were collected by participants during their visit to each institution and were immediately preserved at −20 °C or −80 °C before being transferred to Craif, Inc. for analysis. EVs were isolated from 3 mL of urine samples using Total Exosome Isolation Reagent (from urine) (Thermo Fisher Scientific, Waltham, MA, USA), and total RNA was extracted using the MagMAX mirVana Total RNA Isolation Kit (Thermo Fisher Scientific) according to the manufacturer’s instructions, as previously described^12^.

### Small RNA Sequencing

cDNA libraries were prepared using the QIAseq miRNA Library Kit (QIAGEN, Hilden, Germany) and measured using the Qubit™ dsDNA HS Assay Kit (Thermo Fisher Scientific). Libraries were pooled at equimolar concentrations and sequenced on a NextSeq 550 System (Illumina, San Diego, CA, USA) with single-end reads of 75 nucleotides according to the manufacturer’s instructions. Sequence data were processed as previously described to generate miRNA count profiles^12^.

### Data Preprocessing

A minimum threshold of 10,000 total reads per sample was applied to exclude low-quality or poorly sequenced libraries, as low-depth samples are prone to technical noise and unreliable quantification in urinary miRNA-seq data. miRNAs were retained if they were expressed at a raw count >2 in at least 50% of the samples. Normalization was performed using DESeq2 size factor estimation^14^, followed by log_2_ transformation with a pseudocount of one added to handle zero counts. Differentially expressed miRNAs were identified separately for patients with RCC, PCa, or UC versus healthy controls using the DESeq2^14^ framework (via PyDESeq2). Differential expression was considered significant at adjusted *P* < 0.05 (Benjamini–Hochberg correction) and absolute log_2_ fold change >1.

### Diagnostic Model Development

Cancer-specific diagnostic models were constructed using L2-penalized logistic regression within a nested cross-validation framework^15^ (outer: 5-fold; inner: 3-fold). Feature selection was performed independently within each outer fold using bootstrap-based importance followed by greedy forward selection, retaining the smallest feature set within 0.5% of the maximum cross-validated area under the curve (AUC). The regularization strength was optimized using Optuna’s Bayesian search^16^. Model performance was assessed using aggregated cross-validation predictions and by computing the AUC, sensitivity, specificity, accuracy, precision, recall, and F1 score at both the default and the Youden index thresholds. Confidence intervals were estimated via bootstrapping.

### Prognostic Modeling and Survival Analysis

To evaluate the predictive potential of urinary miRNA signatures for recurrence-free survival (RFS), we first performed univariate Cox proportional hazards regression analysis for all miRNAs associated with postoperative disease recurrence in the RCC, PCa, and UC cohorts. The top 10 miRNAs ranked by the lowest p values were selected per cancer type. All possible combinations of up to five miRNAs from these top 10 were then evaluated in multivariate Cox models. Optimal combinations were selected based on the concordance index performance and proportional hazards assumption validity.

For each cancer type, we generated leave-one-out cross-validated (LOOCV) linear predictors (i.e., risk scores) from the optimal multivariate Cox model. LOOCV was chosen to minimize overfitting and make full use of the limited sample sizes. These predictions were used to stratify patients into high- vs. low-risk groups based on the median value of the predicted risk. Kaplan– Meier curves were plotted and compared via log-rank tests.

We computed time-dependent receiver operating characteristic (ROC) curves^17^ at 48 months using the nearest-neighbor estimator (NNE method) with a smoothing span of 0. LOOCV-based predictions were used as the marker values. Bootstrap resampling (B = 200) was applied to estimate confidence intervals around the ROC curve and AUC. To assess clinical utility, we performed decision curve analysis (DCA)^18^ by comparing the net benefit across a range of threshold probabilities (0.05–0.4). We computed the area under the net benefit curve (AUNB)^19^ for each Cox model relative to the Treat None and Treat All strategies and visualized the delta AUNB per cancer type.

### Statistical Analysis

All analyses were performed using R (version 4.3.2) and Python (version 3.10). Key R packages included survival for survival analysis and Cox regression, survminer^20^ for visualization of Kaplan– Meier curves and forest plots, survivalROC for time-dependent ROC curves, and the base stats package for statistical tests and p value adjustment. Continuous variables are presented as the mean <u>±</u> standard deviation (SD) or median with interquartile range (IQR), and categorical variables are shown as frequencies with percentages. In Python, the analyses utilized scikit-learn^21^ for logistic regression, nested cross-validation, and performance metrics; optuna for Bayesian hyperparameter optimization; numpy^22^ and pandas^23^ for data manipulation; and ggplot2^24^ and ggpubr (for ggarrange) for visualization. P values were corrected for multiple testing using the Benjamini– Hochberg method (p.adjust in R or statsmodels.stats.multitest.multipletests in Python).

## RESULTS

### Baseline Characteristics

A total of 419 urine samples were prospectively collected, comprising three urological cancer cohorts: RCC (n =104), PCa (n =162), and UC (n =153). After quality control and filtering, 96 RCC (92.3%), 153 PCa (94.44%), and 131 UC (86%) samples remained available for analysis. In this study, 455 healthy subjects were included in the control group (see Methods), and subjects adjusted for demographic characteristics, i.e., age, sex and body mass index (BMI), were established as reference groups for each of the three cancer types. **Fig. 1** shows an outline of the analysis pipeline used in this study.

**Fig. 1.**
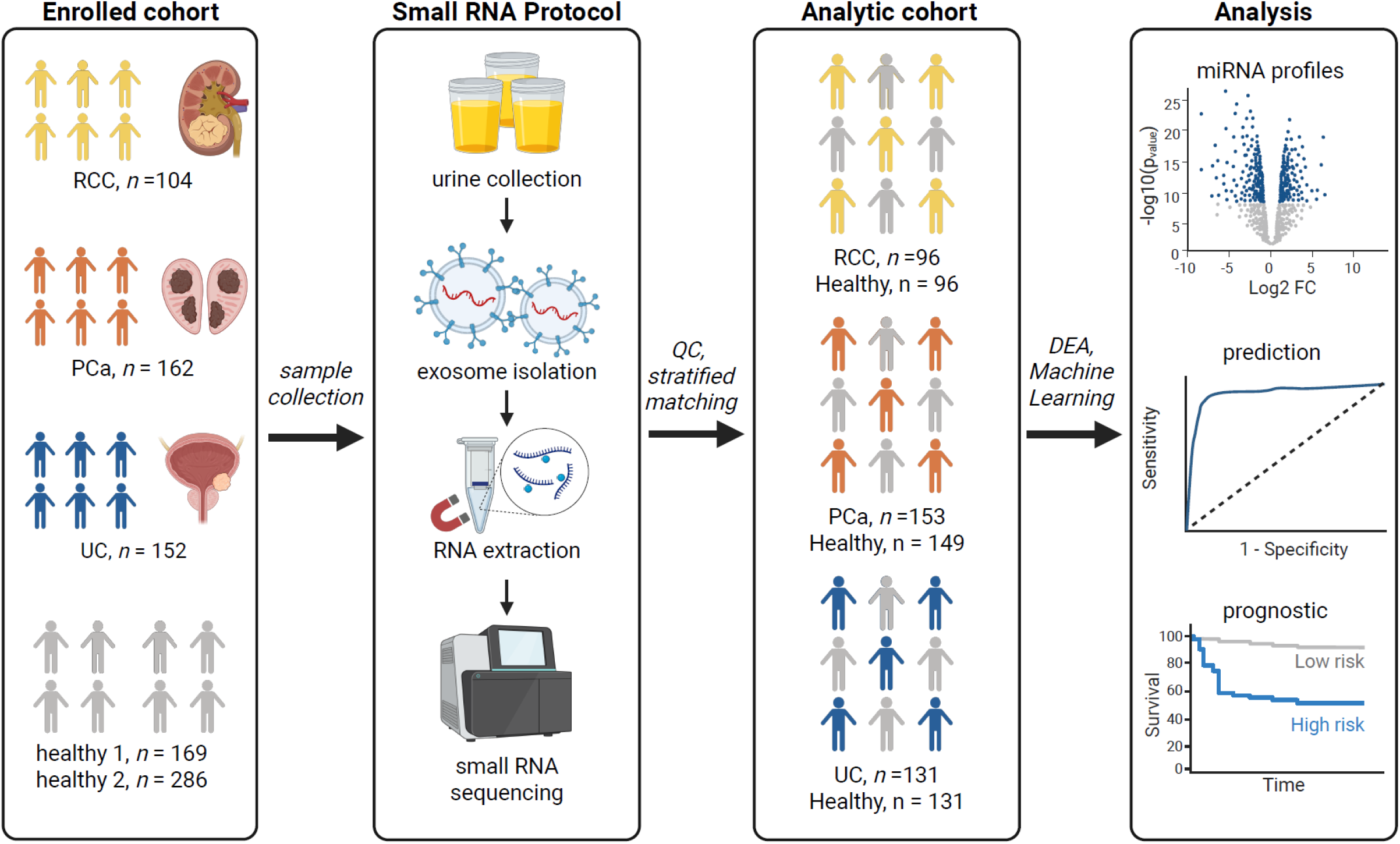
Overview of the study design, including participant enrollment, small RNA sequencing workflow, quality control with stratified matching, and analytic pipeline. Sample sizes for each cancer type (RCC, PCa, UC) and healthy controls are indicated. Downstream analyses included differential expression analysis (DEA), machine learning–based prediction, and survival-based prognostic stratification. QC, quality control; DEA, differential expression analysis.

The median age of patients with UC was 77 years (interquartile range [IQR], 67–87), which was significantly greater than the 66 years (IQR, 59–73) in the control group; the median age of PCa patients was 68 years (IQR, 58–78), which was greater than that of the control group (62 years, IQR, 32–92). On the other hand, patients with RCC were well matched with the control group, with median ages of 64 years (IQR, 48–80) and 64 years (IQR, 60–68), respectively, among the healthy participants. In contrast, the sex distribution and BMI were similar across all the groups (**Table 1**).

**Table 1.** Demographic and clinical characteristics of healthy controls and patients with RCC, PCa, and UC.

| Variable | RCC cohort |  | Pca cohort |  | UC cohort |  |
| --- | --- | --- | --- | --- | --- | --- |
|  | Healthy (n=96) | Cancer (n=96) | Healthy (n=149) | Cancer (n=153) | Healthy (n=131) | Cancer (n=131) |
| **Age** |  |  |  |  |  |  |
| median (IQR) | 63.5 (59.5-67.5) | 63.5 (56.0–72.0) | 62 (32-92) | 68.0 (63.0–73.0) | 66 (59-73) | 77.0 (70.5–81.0) |
| **Sex (%)** |  |  |  |  |  |  |
| female | 32 (33.3) | 25 (26.0) | - | 0 | 23 (17.6) | 28 (21.4) |
| male | 64 (66.7) | 71 (74.0) | 149 (100) | 153 (100) | 108 (82.4) | 103 (78.6) |
| **BMI** |  |  |  |  |  |  |
| median (IQR) | 23.2 (20.2-26.2) | 24.8 (22.5–27.8) | 23.3 (19.3-27.3) | 24.3 (22.5–26.3) | 23.2 (20.2-26.2) | 23.2 (21.3–25.0) |
| **Subgroup (%)** |  |  |  |  |  |  |
| N/A | 96 (100) | - | 149 (100) | 153 (100) | 131 (100) | - |
| ccRCC | - | 79 (82.3) | - | - | - | - |
| non-ccRCC | - | 17 (17.7) | - | - | - | - |
| bUC | - | - | - | - | - | 99 (75.6) |
| UTUC | - | - | - | - | - | 32 (24.4) |
| **AJCC stage (%)** |  |  |  |  |  |  |
| N/A | 96 (100) | - | 149 (100) | - | 131 (100) | - |
| 0/I | - | 78 (81.3) | - | 3 (2.0) | - | 90 (68.7) |
| II | - | 0 (0) | - | 91 (59.5) | - | 9 (6.9) |
| III | - | 12 (12.5) | - | 40 (26.1) | - | 22 (16.8) |
| IV | - | 6 (6.3) | - | 19 (12.4) | - | 10 (7.6) |

Abbreviation: IQR, interquartile range; ccRCC, clear cell RCC; non-ccRCC, non-clear cell RCC; bUC, bladder urothelial carcinoma; UTUC, upper tract urothelial carcinoma.

Pathology features and surveillance data were available for all cancer samples. In RCC, the clear cell type (n = 79, 82%) was predominant, and the nonclear cell type accounted for 18% (17/96 cases). In PCa, all cases were composed of adenocarcinoma, which is the typical histopathology of PCa patients. UC was mainly bladder cancer (n = 99, 76%), and upper urinary tract UC accounted for 24% (32/131 cases) of the entire UC cohort. The disease stages of the three cancer types were as follows: RCC was stage 0/I in 81.3%, stage II in 0%, stage III in 12.5%, and stage IV in 6.3%; for PCa, stage 0/I was 2.0%, stage II was 59.5%, stage III was 26.1%, and stage IV was 12.4%; and for UC, stage 0/I was 68.7%, stage II was 6.9%, stage III was 16.8%, and stage IV was 7.6% (**Table 1**).

### Urinary Extracellular miRNA Profiles in Urological Malignancies

We compared urine-derived extracellular miRNA profiles obtained from patients with RCC, PCa, or UC with those from healthy controls. Compared with the adjusted healthy control group, 24 differentially expressed miRNAs (DEMs) (22 upregulated and 2 downregulated) were identified in RCC (**Supplementary Table 1**), 41 DEMs (36 upregulated and 5 downregulated) were identified in PCa (**Supplementary Table 2**), and 92 DEMs (83 upregulated and 9 downregulated) were identified in UC (**Supplementary Table 3**). In the miRNA expression heatmaps, cancer patients clustered closely within each cancer type, allowing them to distinguish between cancer samples and healthy control samples (**Fig. 2a–c**).

**Fig. 2.**
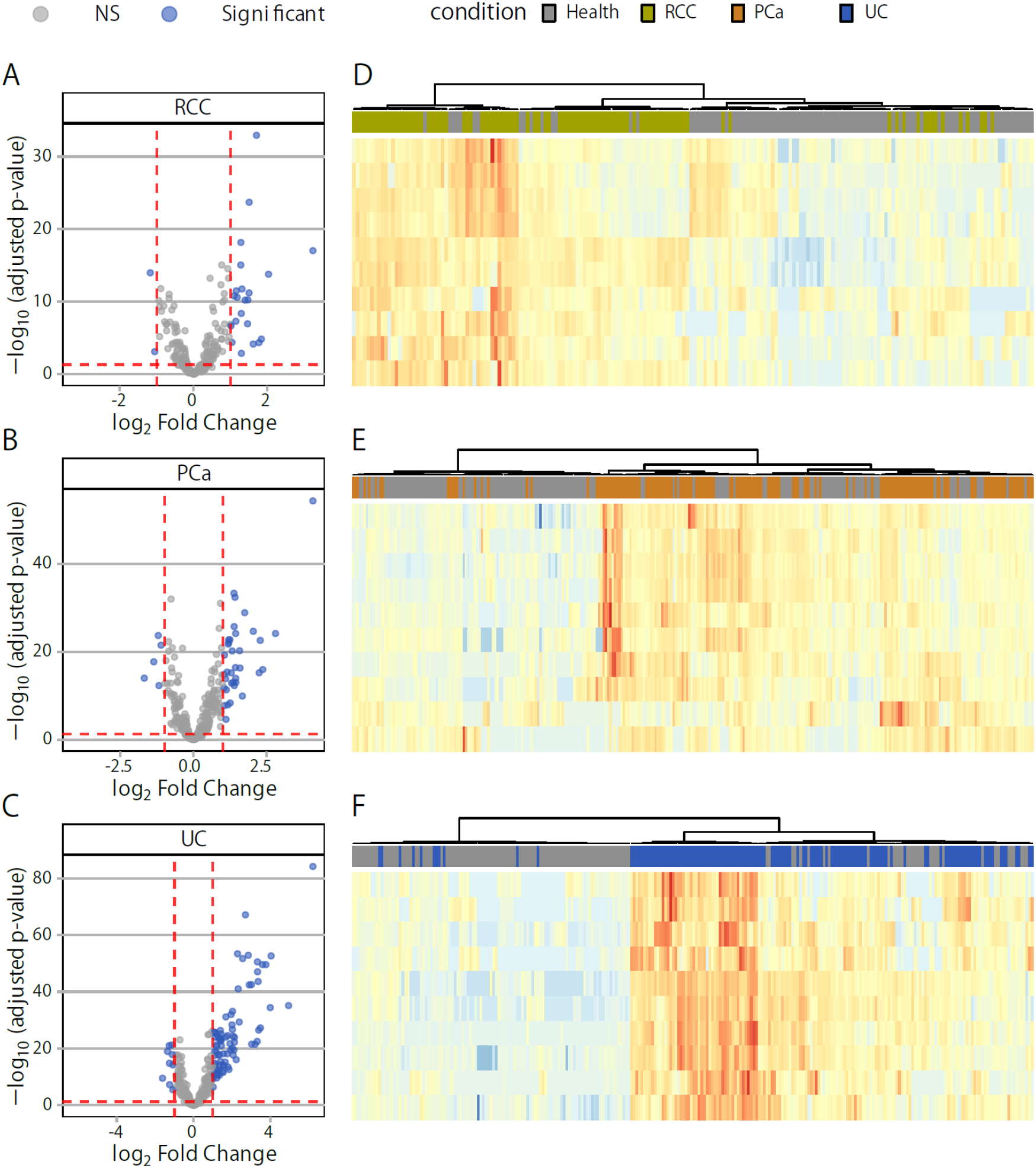
Differentially expressed miRNAs in three urological cancer types. **a-c** Volcano plots and **d-f** heatmaps showing the expression patterns of urinary miRNAs between patients with cancer and healthy individuals in the RCC (**a**,**d**), PCa (**b**,**e**), and UC (**c**,**f**) cohorts. Differentially expressed miRNAs were identified separately for patients with RCC, PCa, or UC versus healthy controls using the DESeq2 framework (via PyDESeq2)^14^.

Hematuria may cause RNA contamination derived from blood, and in our cohort, microscopic or gross hematuria was observed in 10.3% of the RCC cases, 22.0% of the PCa cases, and 52.8% of the UC cases. In contrast, hematuria was observed in less than 10% of the respective healthy control groups. Thus, we excluded samples with hematuria and repeated the differential expression analysis. While the number of significant miRNAs decreased (19 in RCC, 31 in PCa, and 37 in UC), changes in the top-ranked miRNAs persisted (**Supplementary Fig. 1**). These results suggested that the miRNA signatures observed in cancer patients reflect the biological characteristics of the underlying tumor rather than contamination by hematuria in three cancer types.

### Detection Performance for Urological Malignancies

Next, we developed a cancer-specific prediction algorithm using logistic regression classifiers with L2 regularization trained on normalized urinary miRNA expression profiles in each type of cancer. Nested cross-validation was performed to rigorously estimate model performance and optimize hyperparameters. As a result, the ROC curves demonstrated high discriminatory performance for all cancer types, with AUCs of 0.92 (95% CI, 0.88–0.95) for RCC, 0.92 (95% CI, 0.89–0.95) for PCa, and 0.96 (95% CI, 0.94–0.98) for UC (**Fig. 3a**).

**Fig. 3.**
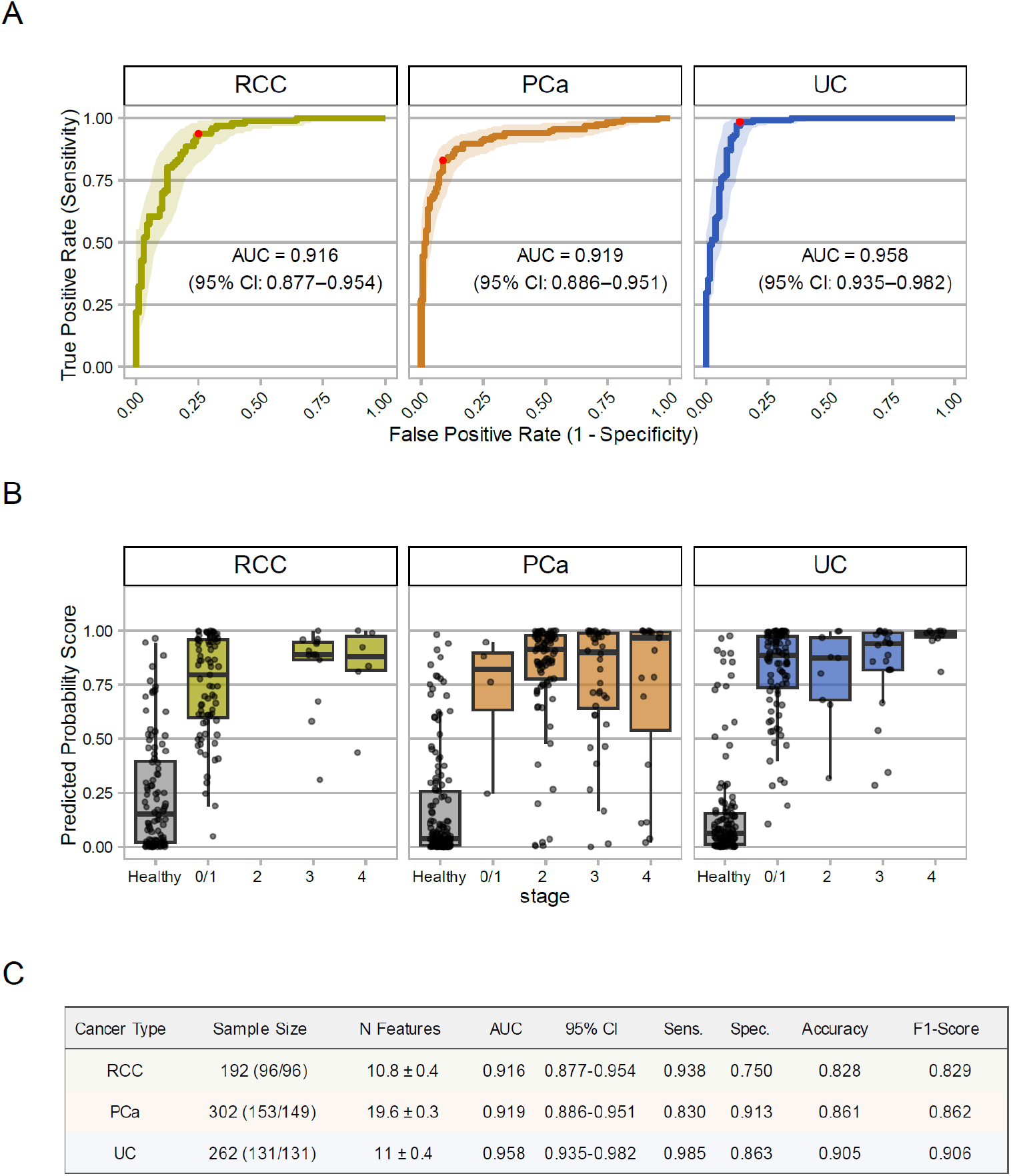
Overview of model performance for urinary miRNA-based classifiers across RCC, PCa, and UC. **a** Receiver operating characteristic curves for each cancer-specific classifier distinguishing healthy individuals from patients with cancer. Shaded areas represent 95% concordance intervals, and red dots indicate the Youden-optimal thresholds. AUC, area under the curve. **b** Predicted probability scores stratified by indicated tumor stage. P values from Wilcoxon rank-sum tests are shown above each group. **c** Summary of classification performance, including sample size, average number of selected features per fold (mean ± SD), AUC with 95% concordance interval, and evaluation metrics at the Youden-optimal threshold: sensitivity, specificity, accuracy, and F1 score.

We then analyzed the model output for each tumor stage. The prediction probability scores obtained from the classifier were stratified based on clinical annotations. The prediction scores for cancer were generally higher than those for the healthy control group and maintained consistently high accuracy across all tumor stages for the three cancer types (**Fig. 3b**). In a subgroup analysis in which changes in predictive scores between histological subtypes were examined, no significant differences were observed between clear cell and nonclear cell types in RCC, however, significant differences were observed between bladder type and upper urinary tract type of UC, with higher predictive accuracy maintained for UTUC (**Supplementary Fig. 2**).

To better understand model behavior, we analyzed the top 20 miRNAs contributing to the classification performance for each type of cancer (**Supplementary Table 4**). The most frequently selected miRNA signatures included hsa-miR-192-5p/215-5p, hsa-miR-27-3p/27b-3p and hsa-miR-574-3p for RCC; hsa-miR-192-5p/215-5p, hsa-miR-2110 and hsa-miR-23a-3p/23b-3p for PCa; and hsa-miR-574-3p, hsa-miR-192-5p/215-5p and hsa-miR-27a-3p/27b-3p for UC. UpSet plots of these microRNA signatures revealed minimal overlap among the three cancer types, revealing that most of the predicted microRNAs are cancer type-specific (**Supplementary Fig. 3**).

Our classification performance is summarized in a comprehensive metrics table, including sample size, area under the curve (AUC) with 95% confidence intervals, and evaluation metrics (sensitivity, specificity, accuracy, and F1 score) computed at the optimal threshold (**Fig. 3c**). Together, confusion matrices generated at a fixed threshold of 0.5 supported these metrics and showed clear separation between the cancer group and the healthy group for the three cancer types (**Supplementary Fig. 4**).

**Fig. 4.**
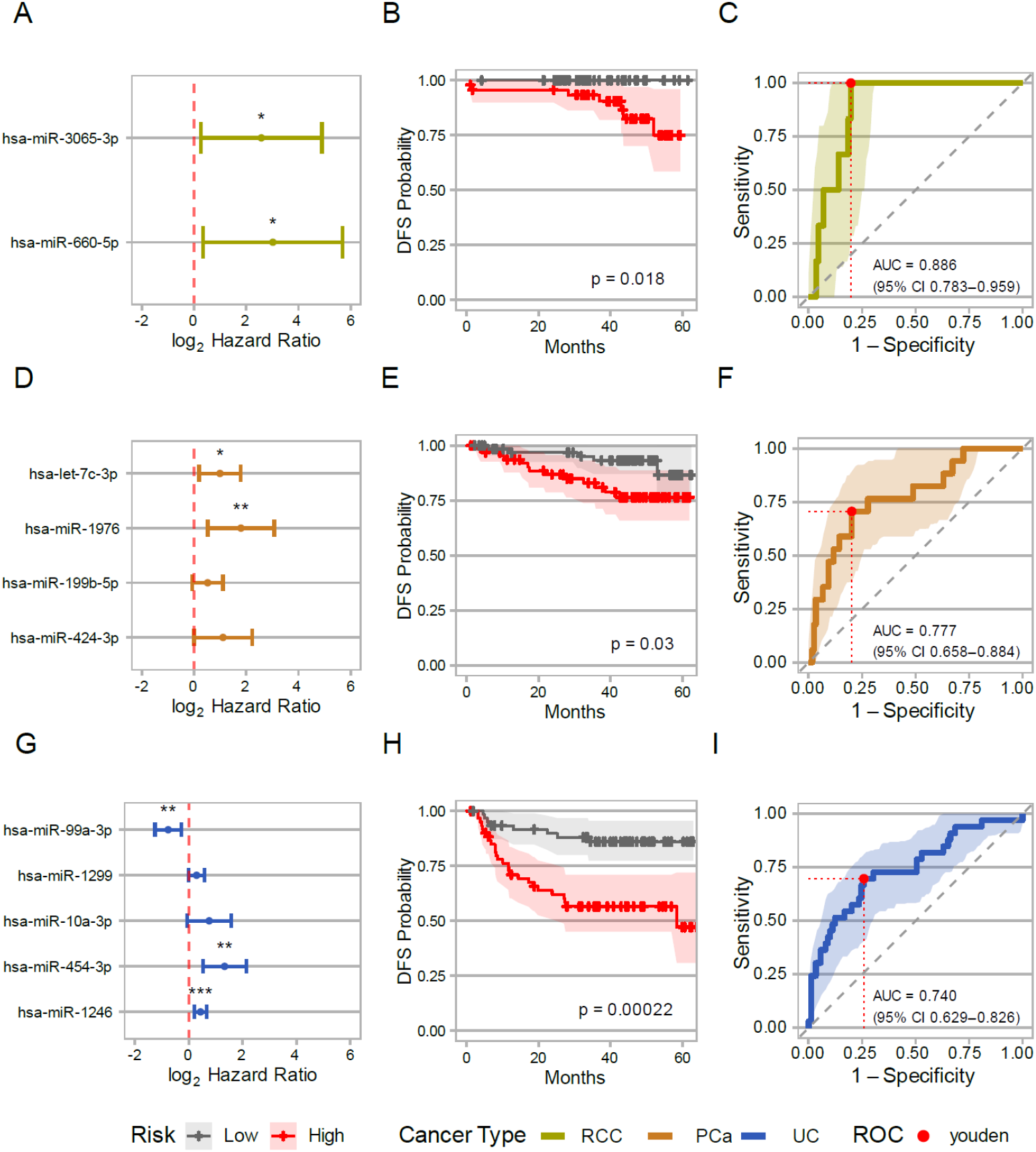
Prognostic performance and survival modeling for urinary miRNA-based classifiers across RCC, PCa, and UC. **a-c** Multivariable Cox regression forest plots showing log_2_ hazard ratios and 95% confidence intervals as forest plots for selected miRNAs included in each cancer-specific model: RCC (**a**), PCa (**b**), and UC (**c**). **d-f** Kaplan–Meier survival curves for each cancer type, stratified by Cox model–derived high-risk and low-risk groups: RCC (**d**), PCa (**e**), and UC (**f**). P values were calculated using the log-rank test. Shaded areas represent 95% confidence intervals. **g-i** Time-dependent receiver operating characteristic (ROC) curves for predicting 4-year recurrence using miRNA-based risk signatures for RCC (**g**), PCa (**h**), and UC (**i**). Shaded regions represent 95% confidence intervals, and red dots indicate Youden-optimal thresholds. The area under the curve (AUC) values and 95% confidence intervals for each cancer type are listed in the corresponding panels.

### Prognostic Performance for Recurrence Prediction

We evaluated the ability of urinary microRNA signatures to predict recurrence-free survival (RFS) in surgically treated patients with RCC, PCa, and UC. During a median follow-up of 43 months (IQR, 36–58), disease recurrence occurred in 7 patients with RCC, 15 with PCa, and 30 with UC.

For each cancer type, we constructed a multivariate Cox model using an optimal panel of up to five miRNAs selected from univariate Cox rankings (see Methods). This process yielded an 11-miRNA prognostic panel across RCC, PCa, and UC (**Fig. 4a–c**), which consistently stratified patients into high- and low-risk groups according to Kaplan–Meier, Cox, and ROC analyses.

Interestingly, several members of this panel have previously been implicated in epithelial– mesenchymal transition (e.g., miR-660-5p, miR-199b-5p, and miR-454-3p), immune evasion (e.g., let-7c-3p and miR-424-3p), and therapeutic resistance (e.g., miR-424-3p and miR-1246)^25–27^. The final models, which were based on the median predicted risk, significantly separated high- and low-risk patients across all cancer types according to Kaplan–Meier analyses **(Fig. 4d–f)**. Discriminative performance was further confirmed by time-dependent ROC analysis at 48 months using leave-one-out cross-validated Cox risk scores, with AUCs of 0.89 (95% CI: 0.78–0.96) for RCC, 0.78 (95% CI: 0.66–0.88) for PCa, and 0.740 (95% CI: 0.629–0.826) for UC (**Fig. 4g–i**).

Finally, decision curve analysis (DCA) was used to evaluate the net clinical benefit of each Cox model. Within a clinically relevant risk threshold range (0.05 to 0.40), all the models demonstrated favorable net benefit profiles. The gain in standardized net benefit (ΔAUNB) was 0.05 for RCC, 0.11 for PCa, and 0.13 for UC (**Supplementary Fig. 5**). Overall, we revealed the potential usefulness of urinary miRNA signatures in recurrence surveillance.

## DISCUSSION

Recent advances in sequencing technology have led to rapid progress in the development of platforms for analyzing miRNAs in exosomes^28^. To date, research on EV-derived miRNAs has focused on blood-derived samples^7,29,30^; however, the invasiveness of blood collection and the risk of infection due to blood contamination remain major challenges.

On the other hand, urine is generally sterile in vivo and does not require complicated postcollection procedures such as hemolysis. Furthermore, urine can be collected by the subject themselves, making it minimally invasive and convenient^12,13^. It is also relatively easy to transport from remote locations, making it an important liquid biopsy material for both subjects and medical aspects. To date, most cancer screening studies based on miRNAs extracted from urinary exosomes have focused on specific miRNAs, and few large-scale prospective studies have comprehensively sequenced miRNA profiles in body fluids. In this study, we revealed two major findings that can be applied to future practice in cancer screening and diagnostics with urinary miRNA signatures.

First, individuals with cancer shed unique EV-derived miRNAs into their bodily fluids, enabling them to be distinguished from healthy controls^31^. Here, we demonstrated that exosomal miRNA profiles in urine can be sequenced, suggesting a novel approach for noninvasive urological cancer screening. Notably, our model can thoroughly screen for cancers from early to advanced stages without overlooking them; therefore, urine-based miRNA profiling may be useful for population-level urological cancer screening and early detection. Hematuria may be a clinical problem due to the contamination of urine with blood-borne miRNAs. However, the miRNA signature observed in this cohort reflects the underlying tumor biology of cancer patients, suggesting that hematuria has a limited impact on urological cancer screening. Interestingly, UpSet analysis of selected features for each cancer type revealed minimal overlap in miRNA panels between cancer types, suggesting the existence of organ-specific exosomal miRNA signatures and the importance of creating machine learning models for individual cancer types.

Second, in addition to cancer screening, this study demonstrated that urinary exosomal miRNAs may be useful for prognostic prediction. Decision curve analysis also demonstrated that urinary exosomal miRNA profiles may be useful for clinical decision-making in posttreatment monitoring across all three cancer types. For the first time, we successfully constructed individualized risk models that effectively stratified RFS using a compact panel of up to five urinary miRNAs for each cancer type, and compared with low-risk patients, patients classified as high risk had significantly worse outcomes according to Kaplan‒Meier curves. Furthermore, at 48 months of follow-up, the time-dependent AUCs of RFS ranged from 0.74 to 0.89 across the three types of cancer, demonstrating high discriminatory power for long-term recurrence. Interestingly, the miRNAs that constitute the prognostic panel have been implicated in processes such as epithelial–mesenchymal transition, immune evasion, and therapeutic resistance^25–27^; therefore, insights into tumor biology may help support the validity of prognosis prediction using our models.

However, our study has several limitations. First, this was from a single center, and a limited number of patients were included. The small number of events, e.g., disease recurrence in a certain cancer subtype, may limit the generalizability of the model. For example, the small number of cases in certain stages and histological subgroups may affect performance estimates and robustness in diagnostic analyses. Also, the small number of recurrence events in each cohort may increase the risk of overfitting in prognostic analyses. Second, this study provides cohort-based cross-sectional analysis results of miRNA profiles in urine collected prior to surgery. Ideally, it is necessary to verify whether the miRNA signature identified as a biomarker is standardized postoperatively or maintains persistently high/low levels and to clarify whether it is useful for monitoring recurrence. Third, biological data to support why specific urinary microRNAs enable the identification of different tumor types or the prediction of recurrence are lacking. Fourth, differences in the storage conditions of urine samples may have introduced variability. Some urine samples from healthy volunteers were stored at −20 °C before transport. Lastly, age distributions slightly differed between UC and PCa cohorts and their respective controls, so residual confounding by age may be a concern.

In summary, our study demonstrated that miRNA profiles derived from urinary EVs could noninvasively distinguish patients with RCC, PCa, or UC from healthy individuals and stratify patients according to their risk of recurrence after treatment. These results position our urine-based assays as promising tools for early detection and surveillance in urologic malignancies. Further validation, i.e., multicenter prospective studies and basic investigations, is needed, advancing such urine testing toward more user-friendly diagnostics and posttreatment management for patients with malignant tumors.

## Supporting information

https://www.dropbox.com/scl/fi/5w2rxdlpivr3puru1j07j/Supplementary-Information.zip?rlkey=q9blnzschddv4vcm0itxe6925&st=wdhq2pm8&dl=0

## Data Availability

All data produced in the present study are available upon reasonable request to the authors.

## Acknowledgments

We would like to thank all our participants and the Omiya City Clinic for contributing to sample collection from healthy individuals. We also thank all laboratory members for their valuable discussions.

## Author Contributions

All the authors reviewed and provided their final approval and agreed to be accountable for all the aspects of this study. Concept and design: YB, SI, MiM, YI, KT, NT, and MO. Sample collection: YK, YY, TT, KM, SM, TK, NT, and MO. Acquisition, analysis, or interpretation of data: YB, SI, AC, HY, YA, and NT. Drafting of the manuscript: YB, AC, HY, and NT. Revision of the manuscript for important intellectual content: All authors. Statistical analysis: AC with HY, YA, MoM, and NT. Administrative, technical, or material support: MoM and YI. Supervision: MoM, MiM, YI, TK, NT and MO.

## Supplementary Information

**Supplementary Fig. 1. Volcano plots of differentially expressed miRNAs between three cancer types and healthy groups after excluding hematuria samples**. Positive log2-fold-change values indicate miRNAs upregulated in cancer samples relative to healthy samples.

**Supplementary Fig. 2. Predicted probability scores from urinary miRNA-based classifiers across different histological subtypes**. Each panel shows model scores for patients in a specific evaluation setting (e.g., held-out test set, postoperative validation, or high-risk assessment), stratified by disease subgroup: clear cell renal cell carcinoma, non-clear cell RCC, bladder urothelial carcinoma, and upper tract urothelial carcinoma. Boxplots represent the distribution of predicted scores, with individual samples overlaid as jittered points. P values for pairwise comparisons were calculated using the Wilcoxon rank-sum test and are displayed above the groups.

**Supplementary Fig. 3. UpSet plot illustrating the overlap and uniqueness of the top-ranked urinary miRNA features among the three cancer types**. The x-axis indicates combinations of cancer types, and the y-axis shows the number of shared miRNAs within each combination.

**Supplementary Fig. 4. Confusion matrices for urinary miRNA-based cancer classifiers across three cancer types**. Faceted heatmaps show binary classification results (cancer vs. healthy) for each cancer type from the nested machine learning test (outer fold) using a fixed decision threshold of 0.5. Each cell reports both the number and percentage of samples for the corresponding predicted vs. actual labels. The color intensity reflects the proportion of samples in each cell. The matrices illustrate the classifier performance across the positive and negative prediction classes.

**Supplementary Fig. 5. Decision curve analysis of Cox regression models for recurrence prediction**. Net benefit curves are shown for RCC, PCa, and UC across threshold probabilities ranging from 5% to 50%. The solid lines represent the Cox model’s net benefit compared with the “Treat None” (dashed) and “Treat All” (dotted) strategies. Shaded ribbons indicate the clinical benefit of the Cox model over the default strategies. ΔAUNB values (differences in area under the net benefit curve) are annotated for each facet.

**Supplementary Table 1.** Differentially expressed 24 miRNAs between RCC and healthy samples. The table reports the mean expression (baseMean), log2-fold change (log2FoldChange), standard error (lfcSE), test statistic (stat), p value, and adjusted p value (padj, Benjamini–Hochberg correction).

**Supplementary Table 2.** Differentially expressed 41 miRNAs between PCa and healthy samples. The table reports the mean expression (baseMean), log2-fold change (log2FoldChange), standard error (lfcSE), test statistic (stat), p value, and adjusted p value (padj, Benjamini–Hochberg correction).

**Supplementary Table 3.** Differentially expressed 92 miRNAs between UC and healthy samples. The table reports the mean expression (baseMean), log2-fold change (log2FoldChange), standard error (lfcSE), test statistic (stat), p value, and adjusted p value (padj, Benjamini–Hochberg correction).

**Supplementary Table 4.** Ranked lists of miRNA feature importance values across RCC, PCa, and UC. For each cancer type, the miRNAs were ranked according to their selection frequency across bootstrap iterations of feature selection. The table includes the full-ranked lists, with the top 20 miRNAs highlighted in Supplementary Fig. 3.

