## Supplementary material for "Urinary miRNA Profiles with Machine Learning for Noninvasive Detection and Prognosis of Urological Malignancies": https://www.dropbox.com/scl/fi/5w2rxdlpivr3puru1j07j/Supplementary-Information.zip?rlkey=q9blnzschddv4vcm0itxe6925&st=wdhq2pm8&dl=0: Supplementary Figs.pdf

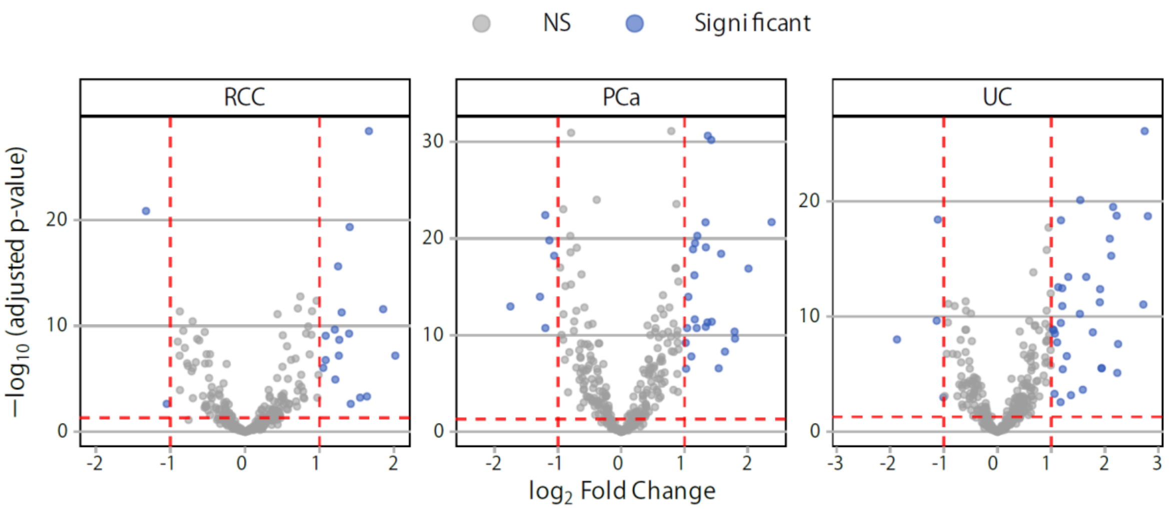


**Supplementary Fig. 1. Volcano plots of differentially expressed miRNAs between three cancer types and healthy groups after excluding hematuria samples.** Positive log2-fold-change values indicate miRNAs upregulated in cancer samples relative to healthy samples.

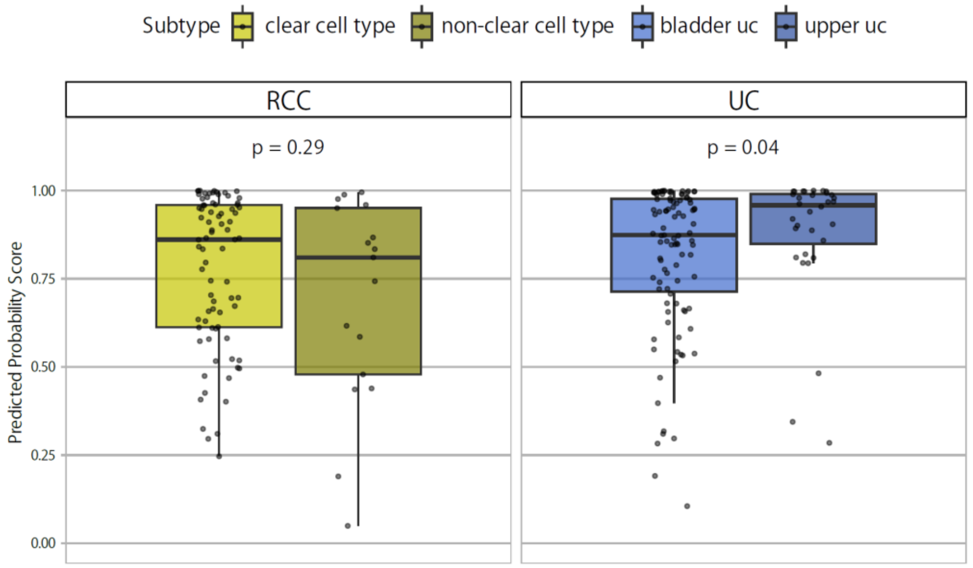


**Supplementary Fig. 2. Predicted probability scores from urinary miRNA-based classifiers across different histological subtypes.** Each panel shows model scores for patients in a specific evaluation setting (e.g., held-out test set, postoperative validation, or high-risk assessment), stratified by disease subgroup: clear cell renal cell carcinoma, non-clear cell RCC, bladder urothelial carcinoma, and upper tract urothelial carcinoma. Boxplots represent the distribution of predicted scores, with individual samples overlaid as jittered points. P values for pairwise comparisons were calculated using the Wilcoxon rank-sum test and are displayed above the groups.

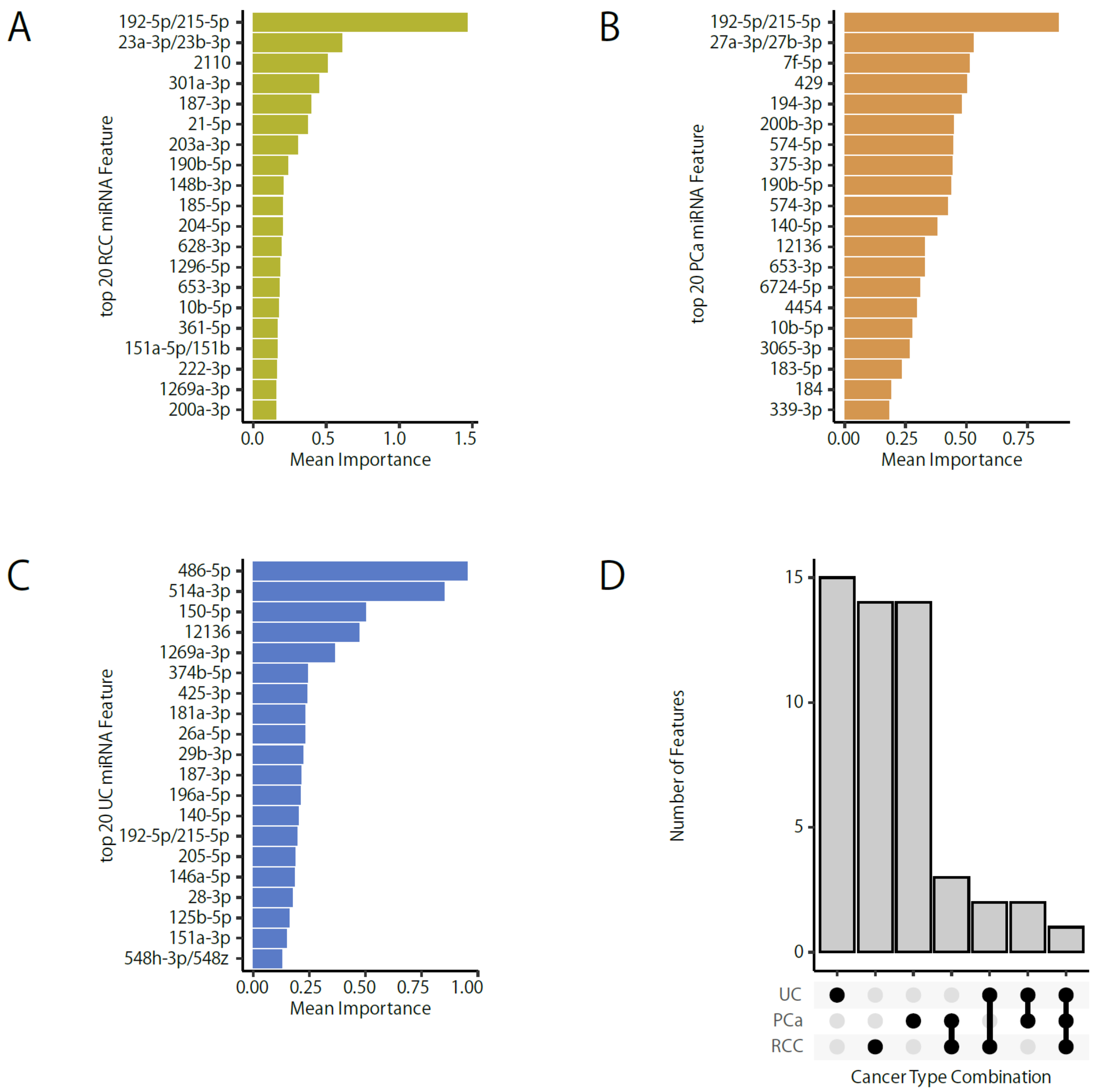


**Supplementary Fig. 3. UpSet plot illustrating the overlap and uniqueness of the top-ranked urinary miRNA features among the three cancer types.** The x-axis indicates combinations of cancer types, and the y-axis shows the number of shared miRNAs within each combination.

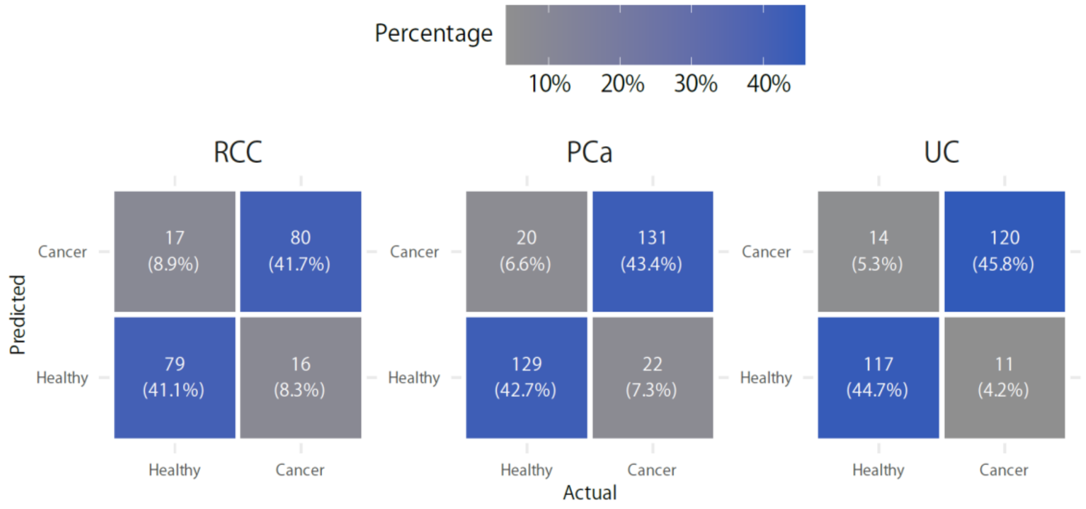


**Supplementary Fig. 4. Confusion matrices for urinary miRNA-based cancer classifiers across three cancer types.** Faceted heatmaps show binary classification results (cancer vs. healthy) for each cancer type from the nested machine learning test (outer fold) using a fixed decision threshold of 0.5. Each cell reports both the number and percentage of samples for the corresponding predicted vs. actual labels. The color intensity reflects the proportion of samples in each cell. The matrices illustrate the classifier performance across the positive and negative prediction classes.

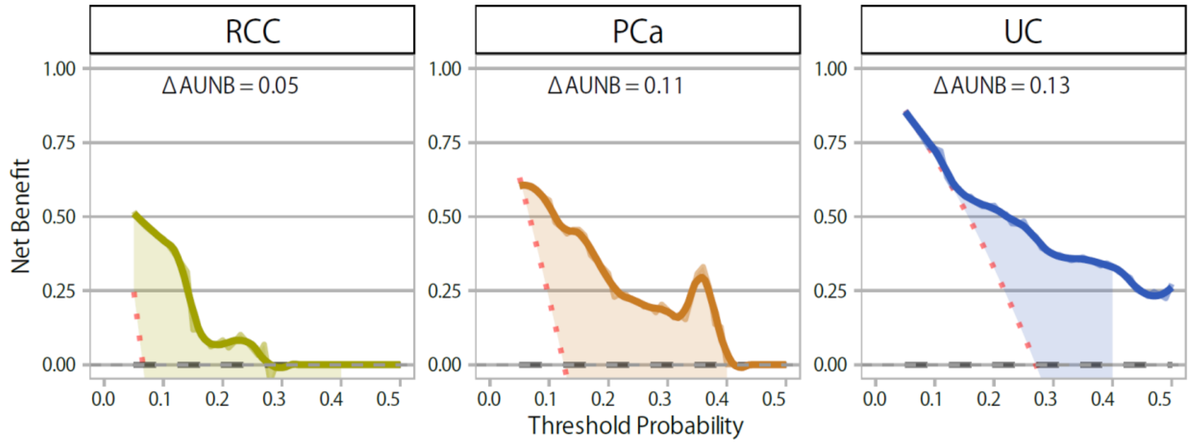


**Supplementary Fig. 5. Decision curve analysis of Cox regression models for recurrence prediction.** Net benefit curves are shown for RCC, PCa, and UC across threshold probabilities ranging from 5% to 50%. The solid lines represent the Cox model's net benefit compared with the "Treat None" (dashed) and "Treat All" (dotted) strategies. Shaded ribbons indicate the clinical benefit of the Cox model over the default strategies. ΔAUNB values (differences in area under the net benefit curve) are annotated for each facet.
